# Safety and long-term performance of the Medtronic 3830 lead in His-bundle vs. Left bundle branch area pacing: A single-center 5-year experience

**DOI:** 10.1101/2024.04.23.24306255

**Authors:** Abdullah Sarkar, Alejandro Sanchez-Nadales, Jose Sleiman, Antonio Lewis-Camargo, Mileydis Alonso, Yelenis Seijo, Andres Sanchez-Nadales, John Bibawy, Marcelo Helguera, Sergio L. Pinski, Jose L. Baez-Escudero

## Abstract

**BACKGROUND:** The short-term safety, feasibility, and performance of the Medtronic SelectSecure 3830-69 cm pacing lead for conduction system pacing (CSP) has been reported; however, its longer-term performance is not well established.

**OBJECTIVE:** The purpose of this study is to examine the long-term performance of the 3830 lead for His Bundle Pacing (HBP) and Left Bundle Branch Area Pacing (LBBAP).

**METHODS:** We retrospectively reviewed all cases of CSP performed with the Medtronic SelectSecure 3830-69 cm pacing lead at Cleveland Clinic Florida between May 2016 and October 2021.

**RESULTS:** Of 515 attempts, HBP achieved an 85% success rate (340 cases), while LBBAP demonstrated a higher success rate of 97.4% (150 cases). The mean follow-up was 28 months for HBP and 14 months for LBBAP, with patient ages averaging 75 and 77 years, respectively. Only 7% of the cohort had an ejection fraction below 50%. The primary indications for HBP were sick sinus syndrome (35.5%), atrioventricular block (35.2%), cardiac resynchronization therapy (10%), and refractory atrial fibrillation (18.8%), with similar distributions for LBBAP. The HBP group’s capture threshold at implant was 1.3 ± 0.8 V at 0.8 ± 0.2 ms, which significantly increased at chronic follow-up to 1.68 ± 1.3 V at 0.7 ± 0.3 ms (p <0.001), whereas the LBBAP group’s capture threshold remained stable from 0.8 ± 0.5 V at 0.5 ± 0.3 ms to to 0.9 ± 0.5 V at 0.5 ± 0.3 ms, (p= 0.35). Lead revisions were more common in the HBP group (50 cases) than in the LBBAP group (5 cases), with exit block rates of 11.7% and 3%, respectively.

**CONCLUSION:** Using the 3830 lead for HBP can result in significantly elevated thresholds, loss of His-bundle capture, and frequent lead revision rates at long-term follow-up. These issues are less commonly seen when the lead is used for LBBAP.

## Introduction

Conduction system pacing (CSP) has emerged as an effective therapeutic strategy for patients with conduction disease. Several advantages to conventional pacing have been reported and include, improvement in cardiac function, reduced QRS duration, and a lower incidence of complications (1–3). Despite the increasing interest and two decades of experience in CSP, there is a dearth of long-term, large-scale, or randomized controlled trials (RCTs), and as such, our understanding of CSP’s strengths and weaknesses remain limited (4).

The reports from current literature on His Bundle Pacing’s (HBP) mechanisms and advantages, highlight the potential to prevent pacing-induced cardiomyopathy and to restore physiological inter-ventricular activation (5,6). However, despite these advancements, HBP has several drawbacks including high thresholds, small sensed R waves, extended fluoroscopy times, and higher rates of lead revision (7,8). CSP later evolved to include Left Bundle Branch Area Pacing (LBBAP) and was noted to have higher success rates, improved R wave amplitudes, and lower pacing thresholds, fluoroscopic times, and procedural times (9,10). Of note, the vast majority of CSP has been performed with the Medtronic Select-Secure 3830 lumenless lead.

We sought to conduct a retrospective analysis of the CSP experience at our center. The operators involved were early adopters of this pacing technique. This study aims to report on long-term outcomes and safety associated with both His Bundle Pacing (HBP) and Left Bundle Branch Area Pacing (LBBAP) using the above mentioned pacing lead.

## Methods

### Data Source, Study Population and Measured Outcomes

Our registry and study protocol were approved by our institutional review board. This retrospective observational study was conducted on all consecutive patients who underwent implantation with the Medtronic SelectSecure 3830-69 cm leads (Medtronic Inc., Minneapolis, MN) for standard indications in accordance with the American College of Cardiology/American Heart Association/Heart Rhythm Society guidelines (11), at Cleveland Clinic Florida – Weston, between May 2016 and October 2021.

The primary endpoint was lead performance data, including sensing and capture thresholds for both HBP and LBBAP. Safety endpoints included system-related complications: infections, lead dislodgment, increased capture threshold (>2.5 V at 1 ms at follow-up or > 1 V from implant), premature battery depletion, and need for lead revision. We also measured the specific left ventricular activation time (LVAT) in HBP and LBBAP patients who were pacemaker dependent **(Supplementary Table 1)**, as previously described (12).

### Implant Procedure

The implantation of all devices involved the SelectSecure 3830-69 cm lead and was conducted by four experienced proceduralists. During the process, an intracardiac electrogram (EGM) from the lead tip and a 12-lead surface electrocardiogram (ECG) were concurrently recorded using a GE CardioLab EP Recording System. The techniques for HBP and LBBAP involved the trans-ventricular-septal approach at the basal ventricular septum. A fixed curve sheath (C315 HIS, Medtronic Inc., Minneapolis, MN) was utilized to deliver the lead. The lead’s tip electrode served for unipolar pacing and mapping the target area during implantation.

For HBP, the procedure aimed to identify the distal His potential, ensuring no atrial electrogram was present before securing the lead. Successful His-bundle capture, either selective (S) or nonselective (NS), indicated a successful procedure, as previously defined (13). If unsuccessful after three attempts, the lead was repositioned for LBBAP or deep myocardial septal pacing in the RV.

LBBAP followed the method outlined by Padala et al. (14). This involved inserting the sheath into the right ventricle, advancing the pacing lead, and applying pacing at 2.0 V/0.5 ms. Successful LBBAP was indicated by a specific QRS morphology in the ECG, characterized by LBBB morphology with a “W” waveform notch at the nadir of the QRS waveform with a progression in ECG morphology until an R’ appears in lead V1. Upon achieving this, the lead was secured with 4–5 clockwise rotations. A high-quality fluoroscopic radiograph was used to confirm the lead’s position towards the septum in the left anterior oblique (LAO) 30° projection, followed by screwing the lead towards the left side of the septum in the right anterior oblique (RAO) 30° projection. If this was not achieved after three attempts, the lead was placed in the right ventricular septum and regarded as left ventricular septal capture (LVSP). In some cases, two sets of 3830 leads and C315 HIS delivery sheaths were used to optimize the pacing site. The first lead marked the His bundle region, while the second targeted the left bundle branch area. Once LBBAP was satisfactory, the first lead was repositioned to the right atrium. If this was not achieved, L

Backup RV pacing leads were implanted in certain patients based on the operator’s judgment, especially early on when experience was limited, using a cardiac resynchronization therapy pacemaker (CRT-P) generator. In these cases, the HBP-RV offset was set at 80ms, or RV output was programmed to subthreshold in non-dependent patients. This strategy aimed to prioritize conduction system pacing while conserving battery life. Initially, HBP lead output was typically set at 3.5 V at 1 ms or twice the His-bundle capture threshold, with auto-capture algorithms disabled.

### Statistical Analysis

The baseline and clinical characteristics of the patients were summarized using descriptive statistics, which included means with standard deviations for continuous variables and counts (percentages) for categorical variables. Univariable analysis was used to compare the characteristics between HBP and LBBAP patients. The independent samples t-test or Mann-Whitney test was used for continuous variables, and chi-square or Fisher’s exact tests were used for categorical variables as appropriate depending on the distribution of the data. A p-value of <0.05 was considered statistically significant. Statistical analyses were performed using SPSS Statistics Version 25.0 (SPSS Inc, Chicago, IL).

## Results

CSP was initially attempted in 515 patients **(Figure 1)**. However, it was unsuccessful in 25 cases. Specifically, 21 patients failed to achieve HBP and were subsequently transitioned to traditional RV pacing. Additionally, 4 patients failed both HBP and LBBAP and also transitioned to RV pacing. These 25 cases, where CSP was not achieved, were excluded from our final analysis. Among the patients included in our complete study cohort, 35 patients initially failed HBP but were successfully transitioned to LBBAP. Overall, our experience achieved a successful CSP implantation rate of 95.1%. When breaking down the success rates, HBP had a 85% success rate, while LBBAP had a 97.4% success rate, (p < 0.001).

**Figure 1:**
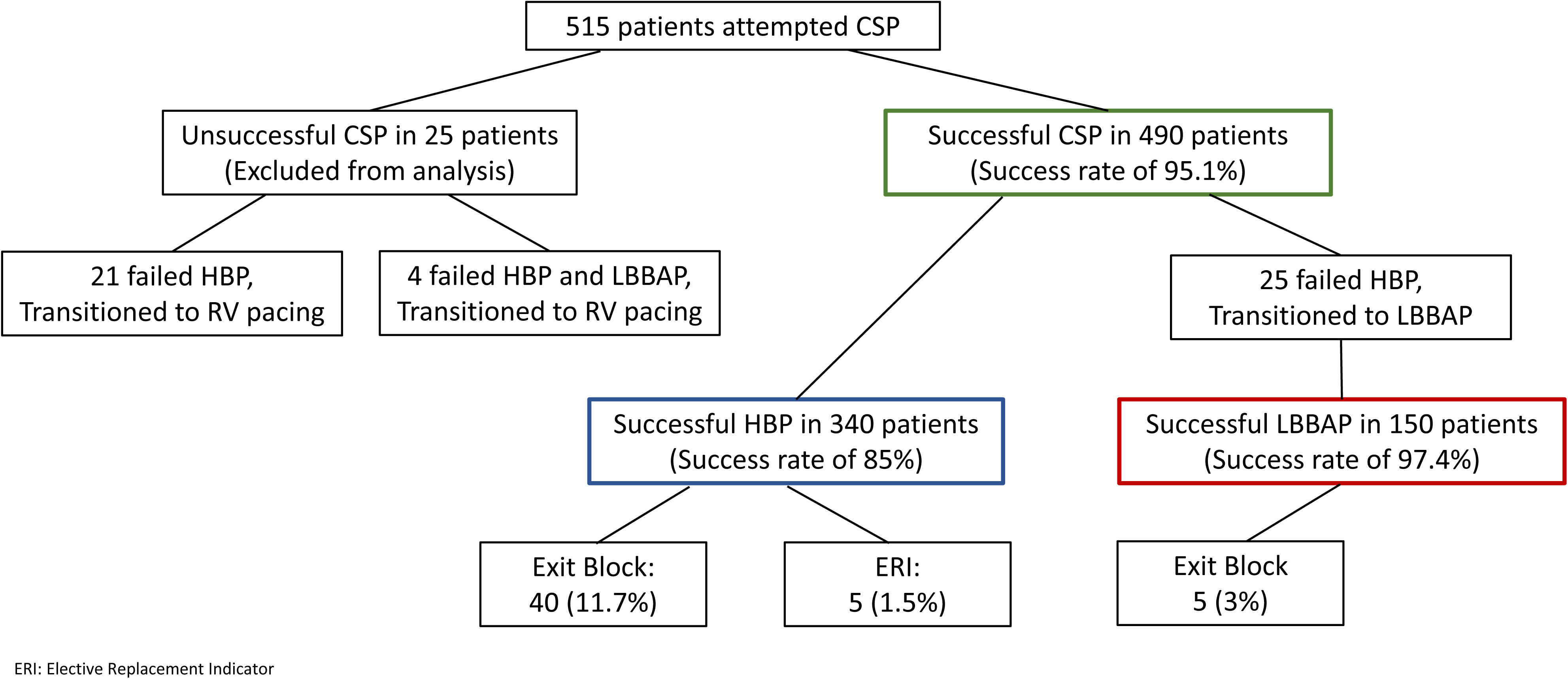
Flowchart of patients that underwent CSP.

### Baseline Characteristics

Our final analysis includes a total of 490 cases with 340 patients in the HBP group and 150 in the LBBAP group. The baseline characteristics and prevalence of several comorbidities are summarized in **Table 1**. The groups were similar in age and gender. The LBBAP group had a higher incidence of congestive heart failure (CHF) (52.6% versus 32.6% in the HBP group, p <0.001) and cardiac amyloidosis (CA) (5.3% versus 1.7% in the HBP group, p= 0.02). In terms of indications, sinus node dysfunction was more prevalent in the HBP group (35.5%) compared to the LBBAP group (24%), (p = 0.11). High degree AV Block was slightly more common in the LBBAP group (46%) than in the HBP group (35.2%), (p = 0.25) The proportion of patients with more than one indication was also comparable between the two groups (23.5% in the HBP group versus 22% in the LBBAP group, p=0.71) **(Figure 2).**

**Figure 2:**
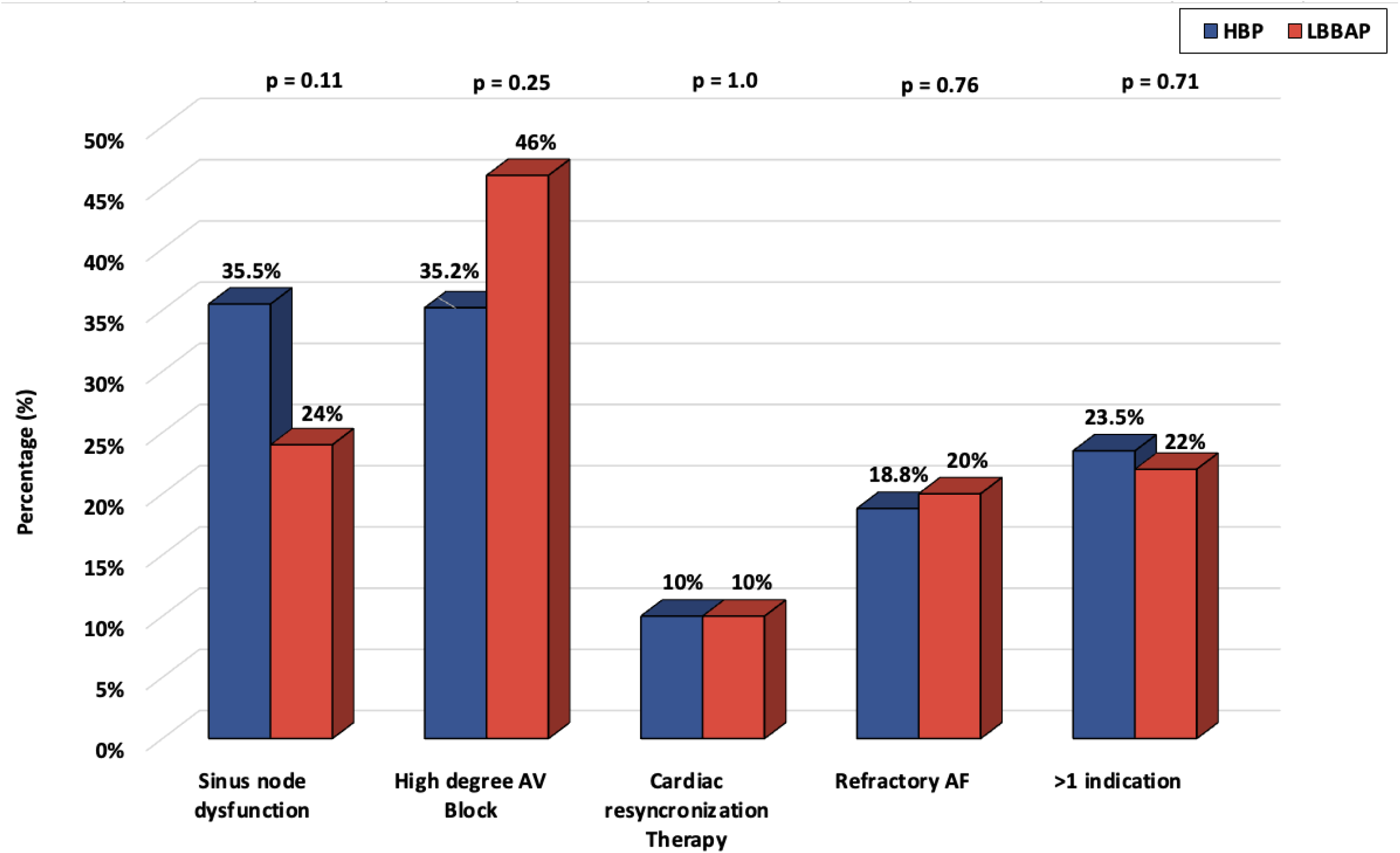
Medical indication for device implantation.

**Table 1:** Baseline Characteristics.

| Variables | HBP (n= 340) | LBBAP (n= 150) | p-value |
| --- | --- | --- | --- |
| **Demographics** |  |  |  |
| Age at implant (years) | 75 ± 9.9 | 77 ± 10 | 0.03 |
| Male Sex | 206 (60.5%) | 94 (62.6%) | 0.66 |
| **Comorbidities** |  |  |  |
| Congestive Heart Failure | 111 (32.6%) | 79 (52.6%) | < 0.001 |
| AF/Flutter | 206 (60.5%) | 103 (68.6%) | 0.108 |
| Hypertension | 233 (68.5%) | 112 (74.6%) | 0.17 |
| Diabetes | 89 (26.1%) | 48 (32%) | 0.18 |
| Cardiac Amyloidosis | 6 (1.7%) | 8 (5.3%) | 0.02 |
| Thyroid Disease | 84 (24.7%) | 43 (28.6%) | 0.35 |
| Creatinine | 1.17 ± 0.82 | 1.17 ± 0.54 | 0.92 |
| **Indications** |  |  |  |
| Sinus node dysfunction | 121 (35.5%) | 36 (24%) | 0.11 |
| High degree AV Block | 120 (35.2%) | 69 (46%) | 0.25 |
| Cardiac Resynchronization Therapy | 34 (10%) | 15 (10%) | 1 |
| Refractory AF | 64 (18.8%) | 30 (20%) | 0.76 |
| >1 indication | 80 (23.5%) | 33 (22%) | 0.71 |
| **Baseline Electrocardiogram** |  |  |  |
| Pre-implant QRS duration (ms) | 113.1 ± 32.6 | 119.5 ± 31.7 | 0.04 |
| Left Bundle Branch Block | 41 (12.05%) | 26 (17.33%) | 0.11 |
| Right Bundle branch Block | 61 (17.9%) | 33 (22%) | 0.3 |
| Paced | 17 (5%) | 10 (6.6%) | 0.47 |

### Procedure Details

The procedure time was longer in the LBBAP group (145 +/- 43 minutes versus 127 +/- 41 minutes in the HBP group, p <0.001). The fluoroscopy time was also slightly longer in the LBBAP group (15 +/- 12 minutes versus 11 +/- 10 minutes in the HBP group, p <0.001) **(Table 2)**.

**Table 2:** Procedure details.

| **Table 2. Procedure Details** |  |  |  |
| --- | --- | --- | --- |
| **Variables** | **HBP (n= 340)** | **LBBAP (n= 150)** | **p-value** |
| Procedure time (min.) | 127 ± 41 | 145 ± 43 | < 0.001 |
| Fluoroscopy time (min.) | 11.3 ± 9.9 | 15 ± 12.4 | < 0.001 |
| Simultaneous AV node ablation | 32 (9.4%) | 21 (14%) | 0.03 |
| Single-Chamber | 18 (5.3%) | 3 (2%) | 0.09 |
| Back-up RV Lead | 21 (6.2%) | 9 (6%) | 0.94 |
| HOT-CRT or LOT-CRT | 16 (4.7%) | 3 (2%) | < 0.001 |

In terms of the type of procedure/device configuration, single-chamber procedures were less common in the LBBAP group (2% versus 5.3% in the HBP group, p <0.001). The LBBAP group had a higher rate of simultaneous AV node ablation (14% versus 9.4% in the HBP group, p=0.03). The use of back-up RV Lead was almost identical in both groups (6% in the LBBAP group versus 6.2% in the HBP group) and was mostly implanted in cases of high degree AV block or simultaneous AV node ablation. The use of His-optimized cardiac resynchronization therapy (HOT-CRT) or Left bundle branch-optimized cardiac resynchronization therapy (LOT-CRT) was less frequent in the LBBAP group (2% versus 4.7% in the HBP group, p <0.001), **(Figure 3)**.

**Figure 3:**
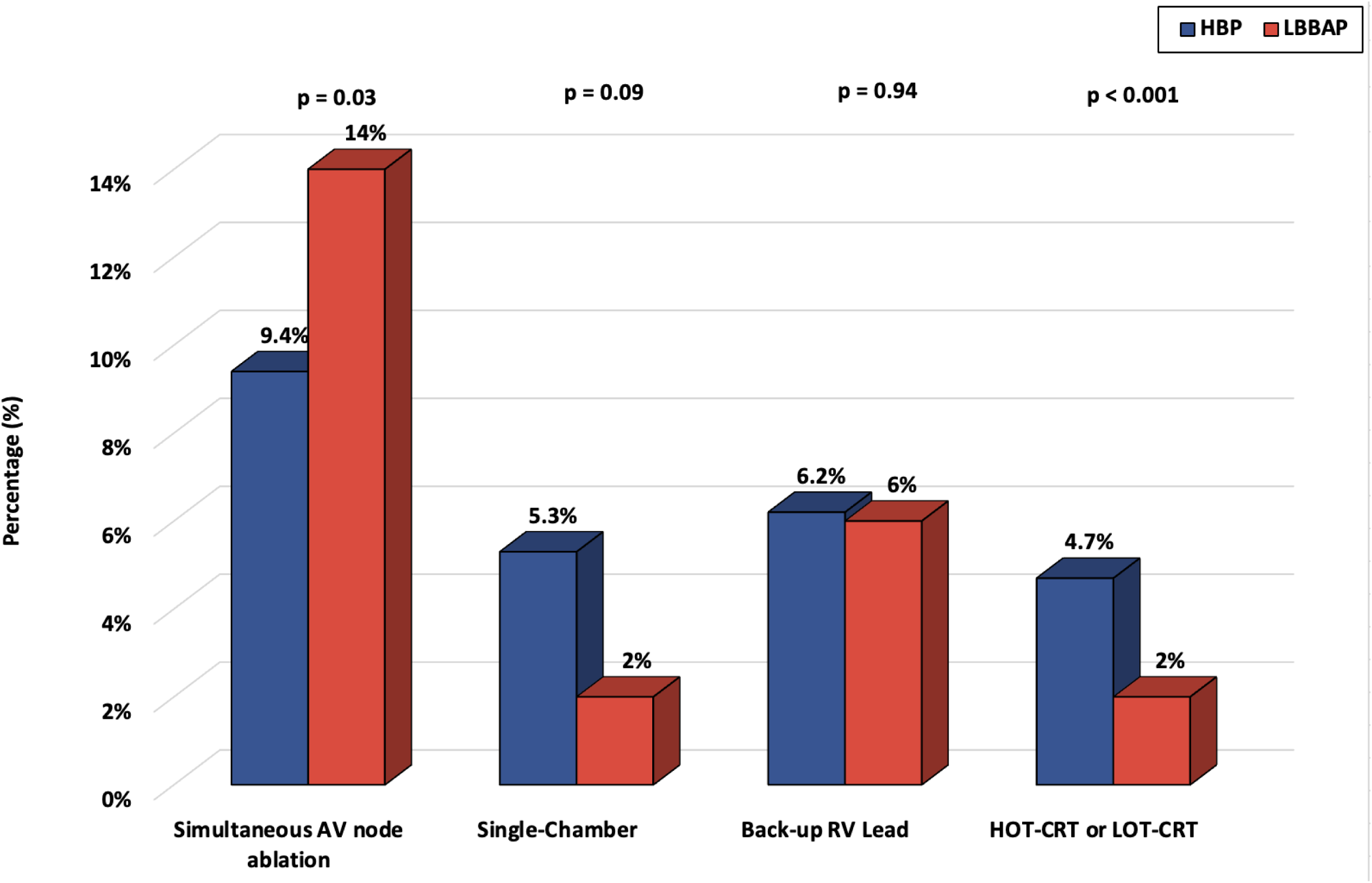
Implant configuration.

### Long-Term Follow-up and Lead Performance

The follow-up period was significantly longer in the HBP group (28 ± 20 vs 14 ± 13 months in the LBBAP group, p<0.001). From the time of device implantation to the latest follow-up, the HBP group experienced an increase in pacing thresholds (from 1.3 ± 0.8 V at 0.8 ± 0.2 ms to 1.68 ± 1.3 V at 0.7 ± 0.3 ms), while the LBBAP group maintained stable thresholds (0.8 ± 0.5 V at 0.5 ± 0.3 ms to 0.9 ± 0.5 V at 0.5 ± 0.3 ms). This resulted in a significant difference in the proportion of patients with a threshold >2.4V at the latest follow-up (18.8% in the HBP group versus 2.6% in the LBBAP group, p<0.001) **(Table 3**, **Figure 4).**

**Figure 4:**
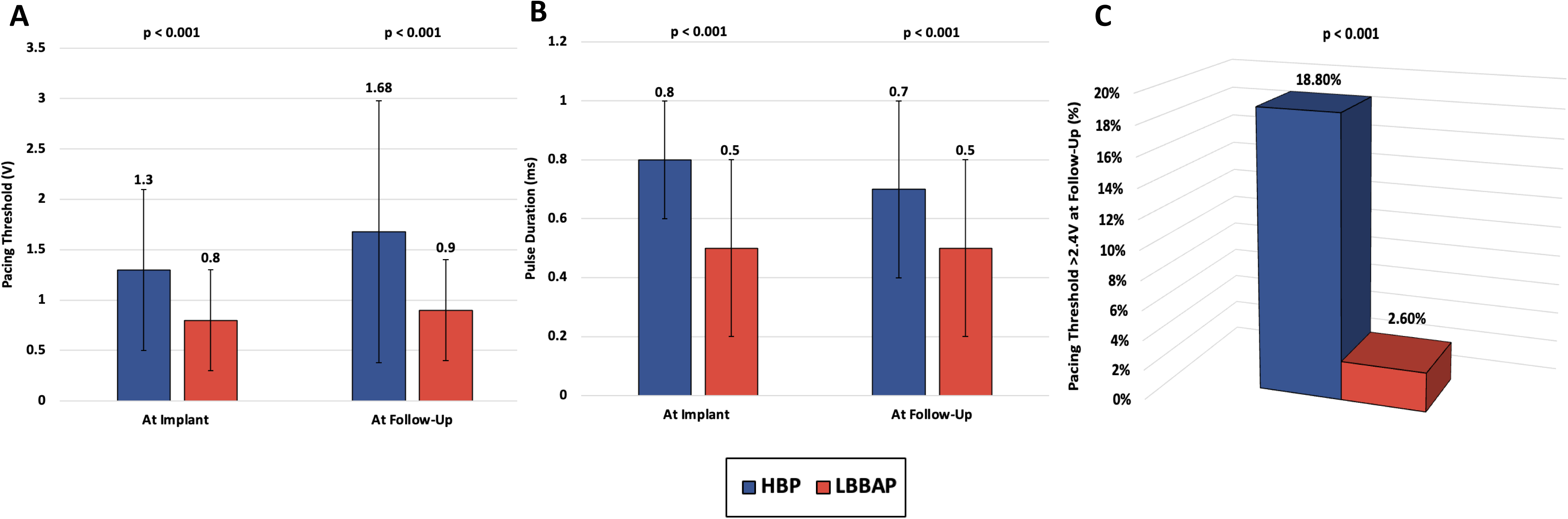
Comparative analysis of pacing parameters (A: Pacing Threshold (V), B: Pulse duration (ms)) measured during implantation and follow-up. C: Percent of patients with pacing threshold >2.4V at follow-up.

**Table 3:** Long-Term Follow-up and Lead Performance.

| **Variables** | **HBP (n= 340)** | **LBBAP (n= 150)** | **p-value** |
| --- | --- | --- | --- |
| Follow up (months) | 28 ± 20 | 14 ± 13 | < 0.001 |
| Loss to Follow-up | 18 (5.3%) | 12 (8%) | < 0.001 |
| Pacemaker dependent | 181 (50%) | 94 (62.6%) | 0.03 |
| **Parameters at Implant** |  |  |  |
| LVEF at implant (%) | 54.4 ± 12 | 53.4 ± 12.9 | 0.46 |
| Post-implant paced QRS (ms) | 133 ± 31 | 138 ± 28 | 0.09 |
| Sensed R wave (mV) | 4.3 ± 2.8 | 9.3 ± 8.7 | < 0.001 |
| Sensed R wave < 2 mV | 46 (13.5%) | 3 (2%) | < 0.001 |
| Impedance (ohms) | 482 ± 125 | 709 ± 165 | < 0.001 |
| Pacing threshold (V) | 1.3 ± 0.8 | 0.8 ± 0.5 | < 0.001 |
| Pulse duration(ms) | 0.8 ± 0.2 | 0.5 ± 0.3 | < 0.001 |
| **Parameters at Long-Term Follow-up** |  |  |  |
| LVEF at follow-up (%) | 53.7 ± 11.4 | 53.7 ± 12.9 | 0.83 |
| Follow-up paced QRS (ms) | 131 ± 36 | 137 ± 31 | 0.118 |
| Sensed R wave (mV) | 5.5 ± 4.5 | 10.7 ± 6.3 | < 0.001 |
| Sensed R wave < 2 mV | 54 (15.8%) | 3 (2%) | < 0.001 |
| Impedance (ohms) | 395 ± 94 | 502 ± 114 | < 0.001 |
| Pacing threshold (V) | 1.68 ± 1.3 | 0.9 ± 0.5 | < 0.001 |
| Pulse duration (ms) | 0.7 ± 0.3 | 0.5 ± 0.3 | < 0.001 |
| Pacing threshold >2.4V | 64 (18.8%) | 4 (2.6%) | < 0.001 |

A subset of patients, 46 (13.5%) initially and 54 (15.8%) at follow-up, had R-wave amplitudes below 2 mV. Conversely, LBBAP cohort, the R-wave amplitudes were higher at implantation, averaging 9.3 ± 8.7 mV, but increased to 10.7 ± 6.3 mV during follow-up (p= 0.007). Low R waves were managed by increasing device sensitivity or switching to a unipolar sensing configuration.

### Safety Endpoints

Within the first 30 days post-implant, two HBP patients developed pneumothorax, which was successfully managed without the need for surgical intervention. Additionally, a pocket hematoma occurred in one HBP patient, treated effectively with conservative medical management. Notably, there were no early reports of device-related infections in either the HBP or LBBAP groups **(Table 4)**.

**Table 4:** Complications within 30-days of implant.

| Variables | HBP (n= 340) | LBBAP (n= 150) | p-value |
| --- | --- | --- | --- |
| **Within 30 Days of Implant** |  |  |  |
| Pneumothorax | 2 (0.6%) | 0% | 0.34 |
| Pocket hematoma | 1 (0.3%) | 0% | 0.5 |
| Device related infections | 0% | 0% | 1 |
| Death < 30 days | 1 (0.3%) | 1 (0.66%) | 0.55 |
| **At Follow-Up** |  |  |  |
| Exit block | 40 (11.7%) | 5 (3%) | < 0.001 |
| Lead extraction | 5 (1.5%) | 0% | < 0.001 |
| Early ERI (<5 years) | 5 (1.5%) | 0% | < 0.001 |

Lead dislodgment within the first month was observed in three cases. Two occurred in the HBP group, with one patient subsequently receiving a re-implantation as HBP and the other as LBBAP. In the LBBAP group it occurred in one patient and underwent re-implantation as LBBAP. Regarding mortality, each group reported a single death within 30 days of implantation. From 2 months to 3 years of follow-up, lead revision was performed in 50 HBP patients and 5 LBBAP patients. The most common reason for of HBP lead revision was a rise in capture thresholds, loss of capture and exit block. Exit block at follow-up occurred in 40 HBP patients (11.7%) compared to 5 LBBAP patients (3%), (p<0.001). ERI were observed in 5 cases of HBP (1.5%), there were no cases of early ERI (<5 years) in the LBBAP group. There were 3 successful lead extractions in the HBP group and no attempts in the LBBAP.

## Discussion

Our single-center study provides insights into the long-term performance and safety of CSP with HBP and LBBAP, utilizing Medtronic SelectSecure 3830-69 cm leads across a substantial cohort of 490 patients. Over an average follow-up of 28 months for the HBP group and 14 months for the LBBAP group, we observed that: 1) HBP capture thresholds significantly increased, with 18.8% of patients exceeding a threshold of 2.5 V; 2) Such an increase was absent in the LBBAP group, which demonstrated lower pacing thresholds and higher R-wave amplitudes; 3) While both techniques proved to be safe, HBP was associated with a lower implantation success rate and higher incidence of lead revision compared to LBBAP.

Our study reports success rates of 85% for HBP (18) and 97.4% for LBBAP (19,20), aligning with the outcomes of other institutions (19,20). The comparatively lower success rates and higher thresholds observed in HBP can be attributed to the anatomical challenges posed by the diminutive and variably positioned His bundle, which measures approximately 2.6 mm in length, 3.7 mm in width, and 1.4 mm in thickness (21), and is surrounded by dense fibrous tissue that is electrically insulating (22). The immediate post-implantation issues such as an acute increase in HBP threshold or loss of capture are often due to suboptimal lead fixation or slight lead displacement. The causes for the delayed and unpredictable rise in HBP threshold over the long term, however, remain unclear.

Our team of implanters have extensive experience with device implantations. Beginning in the latter half of 2018, for patients for whom HBP was not feasible, LBBAP was attempted-initially adopting deep septal pacing techniques. By the middle of 2020, LBBAP had become the predominant method of implantation. In the early phase of our experience, particularly for CSP implants in patients with complete heart block, a back-up RV lead was often placed. Notably, HBP procedures were marked by shorter procedural and fluoroscopy times compared to LBBAP, suggesting a steeper initial learning curve for LBBAP. However, it is implied that with increased experience, the procedural efficiency for LBBAP improved. It’s important to note that HBP was not successful in 25 patients, who were then converted to LBBAP. This conversion contributed to the longer procedural and fluoroscopy times reported for the LBBAP group.

In our study, LBBAP demonstrated a superior lead-myocardial interface, as indicated by higher post-implantation impedance and stability during follow-up. LBBAP was characterized by lower and more consistent pacing thresholds in terms of both voltage and pulse width. Specifically, the mean pacing threshold for the HBP group was 1.3 V at implantation, which significantly increased to 1.68 V (p <0.001) over a median follow-up of 28 months. Notably, 18.8% of patients in this group experienced capture thresholds exceeding 2.5 V which was slightly lower but similar to findings from Teigeler et al. (8) and other centers (9). Conversely, the LBBAP group maintained stable pacing thresholds, with an initial mean of 0.8 V, marginally rising to 0.9 V over a 14-month median follow-up, and only 2% of patients exceeded capture thresholds of 2.5 V. The increasing pacing thresholds observed with HBP highlight a concern for long-term device performance. The higher capture thresholds can lead to faster battery depletion (18) and result in additional procedures, potentially elevating the risk of complications (8). The elevated lead impedances associated with LBBAP suggest a reduced current drain, which may extend battery life comparatively (23).

Our long-term follow-up data reveal that higher capture thresholds in HBP correlate with premature battery depletion, necessitating pulse generator replacement in a significant percentage of patients (18). In fact, within five years, an elective battery replacement indicator (ERI) was activated in 5 HBP patients. In contrast, none of the patients with LBBAP triggered the ERI, however, it should be noted that no LBBAP patients were followed for more than four years **(Supplementary Figure 1).**

In our study, complications were infrequent and typically minor for both groups. We observed no notable differences in the incidence of pneumothorax, pocket hematoma, or device-related infections between the two groups. Each group recorded one death within 30 days post-implantation: an HBP patient died for reasons not determined outside the hospital, and an LBBAP patient passed away due to end-stage cardiomyopathy. It is noteworthy that the latter patient had also undergone implantation of a 4598 lead in the coronary sinus for CRT at the time of the LBBAP procedure.

Supporting our observations, multicenter studies by Padala et al. (19) and Molina-Lerma et al. (24) have documented the safety and dependability of LBBAP, along with its superior pacing performance. Our research not only corroborates these findings but also highlights LBBAP’s consistently lower pacing thresholds. Additional research by Zanon et al. (5) and Hua et al. (25) reinforces the potential of LBBAP as a preferable alternative to HBP and biventricular pacing. Su et al. (26) also acknowledge LBBAP as a safe and feasible option in their single-arm study with follow-up to two years, which our study’s outcomes support but with double the follow-up time.

The initial excitement for HBP to preserve or restore electrical and mechanical synchrony has waned due to various challenges highlighted by our study and experiences at other centers. The pendulum appears to be swinging towards LBBAP, as evidenced by its inclusion in the latest multi-society guidelines. Currently, only LBBAP has been given a Class 2b recommendation as an alternative to RVP for patients with a normal LVEF who are expected to need minimal ventricular pacing (16). Nonetheless, both HBP and LBBAP hold the same recommendation for patients with an LVEF between 36%–50% and anticipated limited ventricular pacing (16).

## Limitations

The field of conduction system pacing is still evolving and thus these findings should be interpreted with caution due to the observational nature of our study and the absence of randomization. The sample size of our study, while substantial compared to current published literature, might limit the statistical power to detect subtle differences between the groups. An inherent learning curve with this emerging technology may account for some of the differences in groups.

Furthermore, our study participants are from a single center, and despite being a tertiary-care referral center with a diverse patient population, the results might not be generalizable. It’s also important to note that patients were lost to follow-up, which could introduce bias into the results. Lastly, the lack of randomization in our study means that there may be unmeasured confounding factors that could have an influence on the results. Further exploration in long-term follow-up studies are needed to validate and expand our understanding of HBP and LBBAP.

## Conclusion

Our retrospective study with patient-level data demonstrates the safety and feasibility of both HBP and LBBAP using Medtronic SelectSecure 3830-69 cm leads. Also, it provides a valuable insight into the comparative performance of HBP and LBBAP in a large cohort of patients with extended follow-up time compared to current literature. LBBAP appears to offer certain advantages over HBP, particularly in terms of a more stable lead-myocardial interface, fewer lead revisions, and potentially longer device longevity.

## Data Availability

Data will be made available upon reasonable request.

## Abbreviations

CSP: Congestive Heart Failure
CSP: Conduction System Pacing
HBP: His Bundle Pacing
LBBAP: Left Bundle Branch Area Pacing
RVP: Right Ventricular Pacing

